# Defining drivers of under-immunisation and vaccine hesitancy in refugee and migrant populations globally to support strategies to strengthen vaccine uptake for COVID-19: a rapid review

**DOI:** 10.1101/2023.03.20.23287477

**Authors:** Anna Deal, Alison F Crawshaw, Jessica Carter, Felicity Knights, Michiyo Iwami, Mohammad Darwish, Rifat Hossain, Palmira Immordino, Kanokporn Kaojaroen, Santino Severoni, Sally Hargreaves

## Abstract

**Background:** Some refugee and migrant populations have been disproportionately impacted by the COVID-19 pandemic, yet evidence suggests lower uptake of COVID-19 vaccines. They are also an under-immunised group for many routine vaccines. We did a rapid review to explore drivers of under-immunisation and vaccine hesitancy among refugee and migrant populations globally to define strategies to strengthen both COVID-19 and routine vaccination uptake.

**Methods:** We collected global literature (01/01/2010 - 05/05/2022) pertaining to drivers of under-immunisation and vaccine hesitancy in refugees and migrants, incorporating all vaccines. We searched MEDLINE, Embase, Global Health PsycINFO and the WHO’s ‘Global Research on COVID-19’ database and grey literature. Qualitative data were analysed thematically to identify drivers of under-immunisation and vaccine hesitancy, then categorised using the ‘Increasing Vaccination Model’.

**Results:** 63 papers were included in this review, reporting data on diverse population groups, including refugees, asylum seekers, labour and undocumented migrants from 22 countries, with six papers reporting on a regional or global scale. Drivers of under-immunisation and vaccine hesitancy pertaining to a wide range of vaccines were covered, including COVID-19 (n=27), HPV (13), measles or MMR (3), influenza (3), tetanus (1), and vaccination in general. We found a range of factors driving under-immunisation and hesitancy in refugee and migrant groups, including unique awareness and access factors that need to be better considered in policy and service delivery. Acceptability of vaccination was often deeply rooted in social and historical context and influenced by personal risk perception.

**Conclusions:** These findings hold direct relevance to current efforts to ensure high levels of global immunisation coverage, key to which is to ensure marginalised refugees and migrant populations are included in national vaccination plans of low-middle- and high-income countries. We found a stark lack of research from low- and middle-income and humanitarian contexts on vaccination in mobile groups, a situation that needs to be urgently rectified to ensure high coverage for COVID-19 and routine vaccinations.

## Introduction

There are an estimated one billion people on the move globally (1 in 7 of the global population), with refugee and migrant populations known to have been disproportionately impacted clinically and socially by the COVID-19 pandemic (1, 2). However, despite their increased risk from infection and potentially adverse outcomes, refugees and migrants have shown lower COVID-19 vaccination uptake in the few countries where this has been measured (3–8). For examples, in a recent study of 465,470 migrants in the UK, uptake of the first dose was reported to be slower across all age groups for migrants compared with the general population. Refugees and older migrants were more likely to have delayed uptake of COVID-19 vaccines and to not have received their second or third dose. Refugees specifically had a higher risk of a delayed second dose (odds ratio 1·75 [95 CI% 1·62–1·88]) and third dose (1·41 [1·31–1·53]) (9). Large country and regional differences in COVID-19 vaccine acceptance rates have been reported among all population groups (10). These datasets are a brief snapshot in time in what is a rapidly evolving field; however, they suggest that innovative strategies are likely needed to improve COVID-19 vaccine access and uptake in these populations in the immediate term as COVID-19 vaccines become more widely available globally (including through the COVAX Facility and the COVAX Humanitarian Buffer), alongside ongoing work to strengthen routine vaccination uptake in refugees and migrants in the longer-term (11) (12, 13).

In recent years, debates around vaccination acceptance and intent have become increasingly complex. The term ‘vaccine hesitancy’ has been defined by the WHO Strategic Advisory Group of Experts on Immunization (SAGE) Working Group on Vaccine Hesitancy as ‘…the delay in acceptance or refusal of vaccination despite availability of vaccination services’ (14); it was in 2019 considered among the top ten threats to global public health (15). Vaccine hesitancy is complex and highly variable in different contexts and over time (14, 16, 17), yet little focus to date has been placed on determinants of vaccine hesitancy in refugee and migrant populations. The ‘3Cs’ model describes vaccine hesitancy as being driven by the ‘3 Cs’: Confidence (importance, safety, and efficacy of vaccines), convenience (access issues dependent on the context, time and specific vaccines being offered) and complacency (perception of low risk and low disease severity) (16, 18). Researchers, however, have stressed the need to find better terms to clearly distinguish between vaccine hesitancy (more focused around personal/psychological influences) and the other determinants of uptake, such as logistical problems and physical barriers to accessing vaccines in order to ensure vaccination initiatives are successful, factors that will be particularly pertinent in refugee and migrant populations (19). The Increasing Vaccination Model, recently adapted by the WHO expert working group to measure behavioural and social drivers of vaccination (BeSD) to support more comprehensive planning and evaluation around vaccine uptake, bring together various models and frameworks into a working model to conceptualise drivers of under-immunisation measuring three domains that influence vaccine uptake: what people think and feel about vaccines; social processes that drive or inhibit vaccination (which both combine to influence individual motivations, or hesitancy, to seek vaccination); and practical factors involved in seeking and receiving vaccination (20–22). The WHO expert working group has now begun the development of globally standardised tools for health policy makers and planners to measure and monitor reasons for under-immunisation in real time, including for COVID-19 (20, 22).

Refugees and migrants may face a range of unique personal, social and physical barriers to accessing health and vaccination services, which may influence vaccine motivation. This may be particularly so among refugees and migrants who are new to the host country, those with precarious immigration status, and those residing in camps and detention facilities who may be excluded from mainstream health and vaccination systems (2, 11), with evidence that they are an under-immunised group for routine vaccinations (4, 11, 23, 24). Drivers of under-immunisation and hesitancy may include difficulty understanding the local healthcare system, language barriers, discrimination or racism, and real, restricted or perceived lack of entitlement to free vaccinations, low trust in health systems, cultural barriers, and/or being unable to afford direct or indirect costs (4, 12, 25–31). One systematic review exploring the role of refugees and migrants in outbreaks of vaccine-preventable diseases in Europe reported a high number of outbreaks among adult and child migrants in temporary refugee and migrant camps, linked to lack of access to mainstream vaccination systems (24). Some refugee and migrant populations will face specific barriers to public health messaging that will impact on vaccine motivation (31–35), with a subsequent impact on vaccination uptake in some communities (33). Uptake data and drivers of under-immunisation are often lacking in these populations, and there may be important differences between refugees and migrants residing in high-income, compared to low- and middle-income settings or in specific humanitarian contexts (including closed camp settings or detention centres), for example, that are yet to be fully elucidated. This suggests that that more research is urgently needed to explore and assess drivers of under-immunisation and vaccine hesitancy in diverse refugee and migrant populations globally in order to define evidence-based solutions to support COVID-19 vaccine roll-out.

We therefore did a rapid review of published and grey literature to explore key drivers of under-immunisation and vaccine hesitancy among diverse refugee and migrant populations in low-middle- and high-income countries and humanitarian settings, to explore and assess country and regional differences, and to document strategies, solutions, and best practices to strengthen vaccination uptake for COVID-19 in the immediate term.

## Methods

We did a rapid review of published and grey literature pertaining to refugee and migrant communities globally. We followed guidelines developed by the Joanna Brigg’s Institute (JBI) for scoping reviews (36), as well as the Preferred Reporting Items for Systematic Reviews and Meta-Analyses extension for Scoping Reviews (PRISMA-ScR) checklist (37).

### Inclusion and exclusion criteria

We collected global published literature pertaining to drivers of under-immunisation and vaccine hesitancy in refugees and migrants for all vaccines including low-skilled labour migrants, asylum seekers, undocumented migrants, migrant health-care workers and others, residing in all low-middle- and high-income countries, and including humanitarian settings.

We defined vaccine hesitancy, as ‘the delay in acceptance or refusal of vaccination despite availability of vaccination services’ (14) (38). Refugees are defined in the Convention and Protocol Relating to the Status of Refugees as “persons outside their countries of origin who are in need of international protection because of feared persecution, or a serious threat to their life, physical integrity or freedom in their country of origin as a result of persecution, armed conflict, violence or serious public disorder.” (see https://www.unhcr.org/3b66c2aa10 and https://www.unhcr.org/master-glossary.html). We defined a migrant as an individual born outside of their current country of residence (see https://publications.iom.int/system/files/pdf/iml_34_glossary.pdf); however, where this information was not available in the literature, first language, nationality or the paper’s own definition of a migrant were used as a proxy, in order to avoid excluding relevant literature. Articles published between January 1, 2010 and April 5, 2022 were eligible for inclusion, with no restrictions on language. Articles focusing on drivers of under-immunisation and vaccine hesitancy in the wider population with results not disaggregated by legal status were excluded. Papers were not excluded based on study type, and opinion pieces, commentaries, guidelines, policy briefs, and review articles were eligible for inclusion where they met our inclusion criteria.

### Search strategy

We searched MEDLINE, Embase, Global Health PsycINFO and the WHO’s Global research on COVID-19 database (https://www.who.int/emergencies/diseases/novel-coronavirus-2019/global-research-on-novel-coronavirus-2019-ncov) for literature pertaining to drivers of under-immunisation and vaccine hesitancy in migrant populations globally. The search strategy used keywords relating to migrant populations, vaccine hesitancy, and barriers to vaccination (Table 1). Subsequently, grey literature sources were searched for, including through the following websites: World Health Organization (WHO), International Organization for Migration (IOM), European Centre for Disease Prevention and Control (ECDC), Doctors of the World, Médecins sans Frontières (MSF), a variety of humanitarian and wider resources and websites (including the British Red Cross Community Engagement Hub, IFRC, IOM, Reliefweb.org, UNICEF), as well as Google and Google Scholar. Experts from key countries and relevant organisations were engaged to gather further literature.

**Table 1.** Search Strategy

| Theme | Keywords |
| --- | --- |
| Migrants | migrant* OR immigrant* OR emigrant* OR foreign* OR asylum* OR asylee* OR refugee* non-citizen* OR citizenship OR nationality OR undocumented OR non-resident* OR expat* OR newcomer* |
| Vaccination | vaccin* or immunis* or immuniz* |
| Drivers of under-immunisation and vaccine hesitancy | (uptake or demand or coverage or utiliz* or utilis*) OR<br>(barrier* or enabl* or facilitat* or motivat* or obstacle* or determinant* or factor* or reason* or challenge*) OR<br>(accept* or comply or complian* or adher* or readiness or intent* or willing*) OR<br>(avoid* or refus* or hesita* or renounc* or reject* or deny or deni* or delay*) OR<br>(confiden* or trust* or fear* or wary or wariness or doubt* or sceptic* or concern* or complacen*) OR<br>(attitude* or perception* or perspective* or view* or belief*) OR<br>(practice* or behaviour*) OR<br>(decision or decision-making) OR<br>(aware* or knowledge* or inform* or understand*) OR<br>(access* or cost* or afford* or navigat* or availab*) |
| Years | 2010 – 2022 |

### Data extraction, synthesis and analysis

Data were extracted by AD, with SH duplicating data extraction in 50% of included papers. Qualitative data were first analysed thematically to identify drivers of under-immunisation and vaccine hesitancy, then categorised using the WHO’s Increasing Vaccination Model (21, 22). Using Microsoft Excel, a standardised form was developed to extract data on the following: Author and year of publication of study, study setting and location, study design, vaccine(s) studied (if concerned with a specific vaccine), number of participants (where relevant), refugee and migrant demographics (country of origin, legal status), age group [e.g., children, adolescents, adults], gender), drivers of vaccine hesitancy, and recommended strategies, solutions, and best practices to strengthen uptake of vaccines.

## Results

Our searches identified 737 records, of which 631 were identified as unique. After title, abstract and full-text screening, 63 papers were identified and included in this review, see Figure 1.

**Figure 1.**
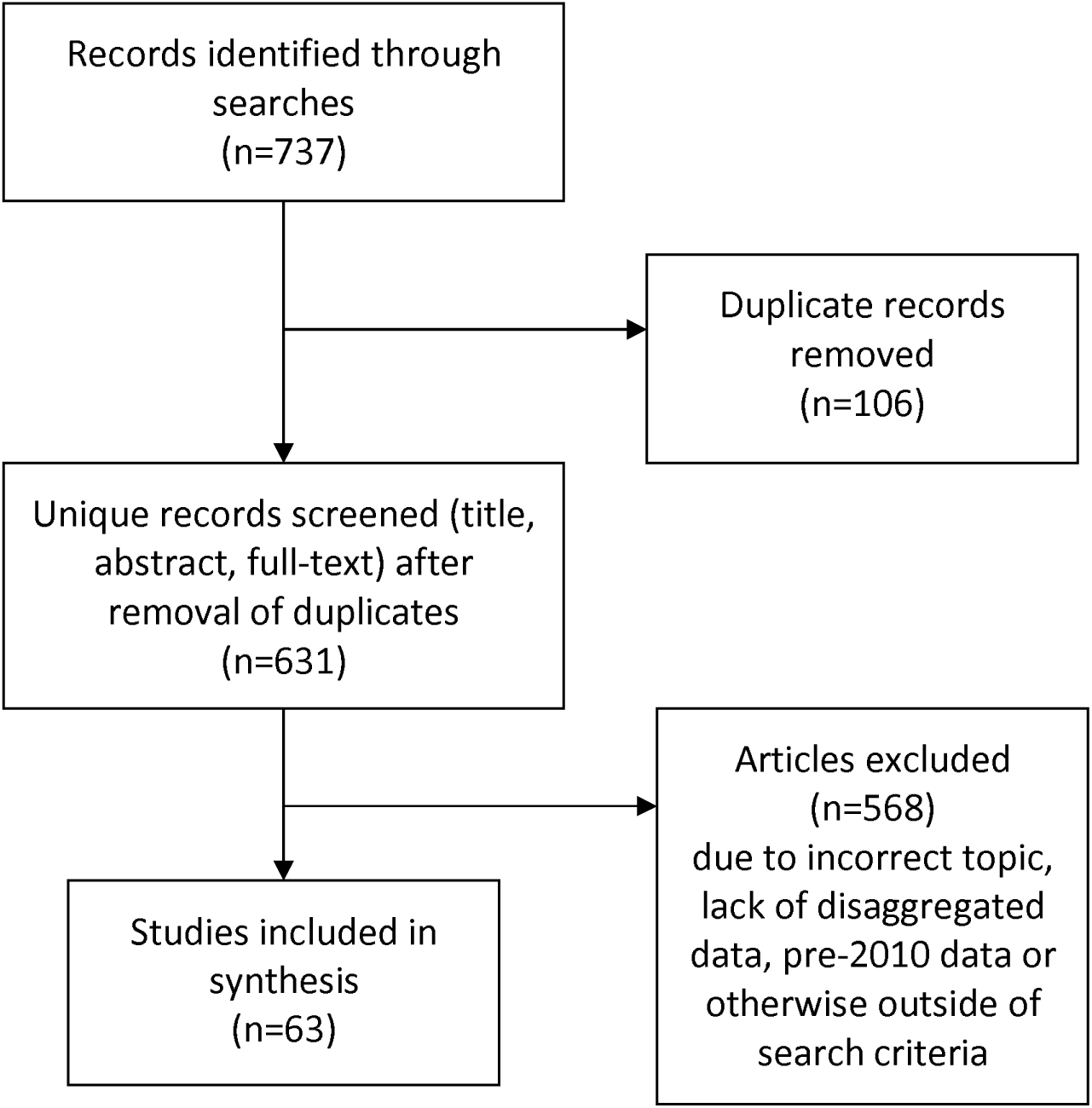
PRISMA diagram of included and excluded studies

Included papers summarised in Table 1 reported data on migrants and refugees from 22 countries Australia, Bangladesh, Canada, Denmark, France, Greece, Hungary, Italy, Japan, Lebanon, Netherlands, Norway, Poland, Qatar, South Korea, Sweden, Switzerland, UAE, USA, UK), with six papers reporting on a regional or global scale (Europe, Africa). Papers covered a wide range of specific vaccines, including COVID-19 vaccines (n=27), HPV vaccines (13), measles or MMR vaccines (3), influenza vaccines (3), and tetanus vaccines (1), with other papers focusing on vaccination in general and/or childhood vaccines. Multiple migrant types, different nationality groups, and contexts were covered in the included literature, including data pertaining to refugees, asylum seekers, labour migrants, and undocumented migrants, in both community (urban and rural) and humanitarian settings.

**Table 1.**
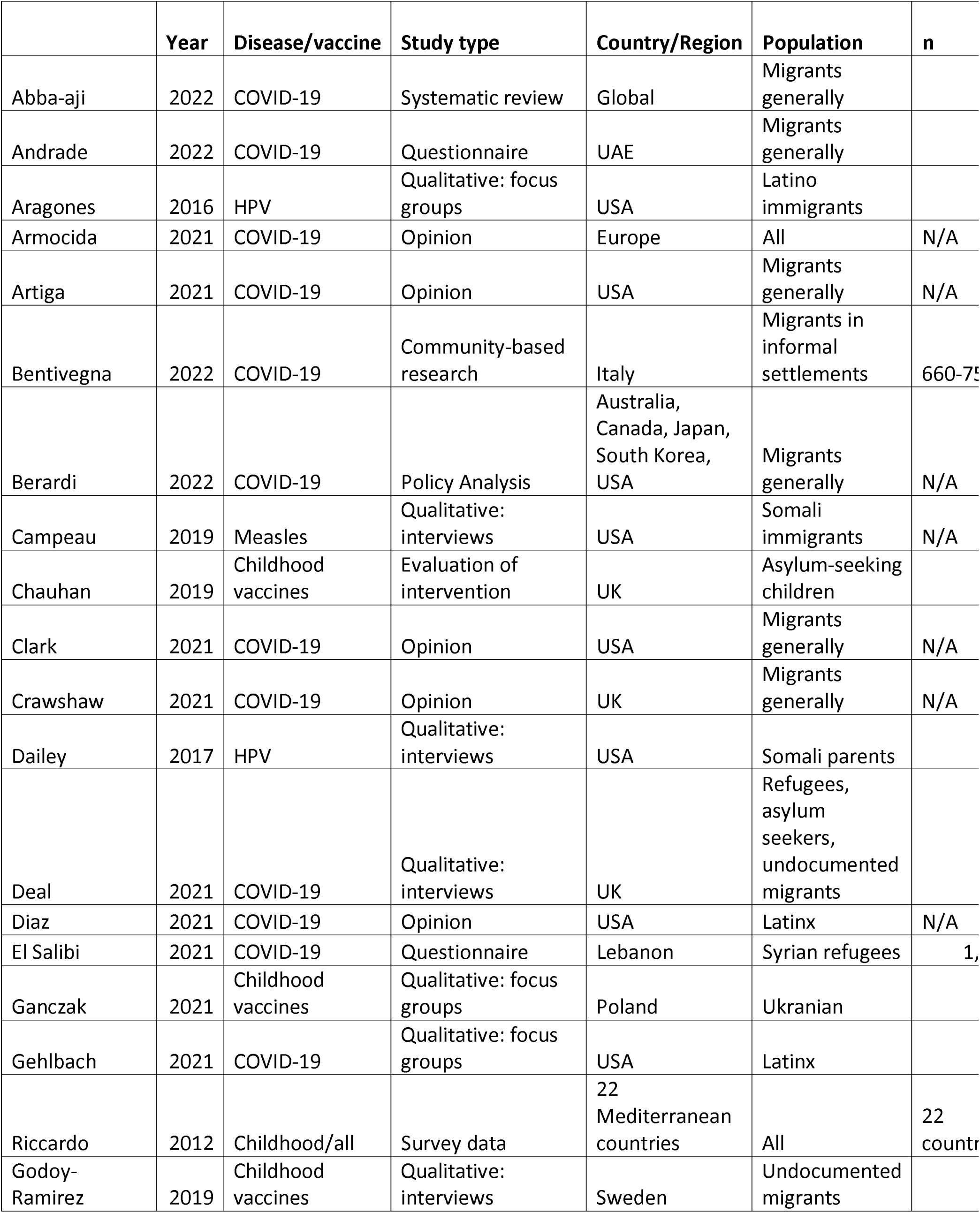
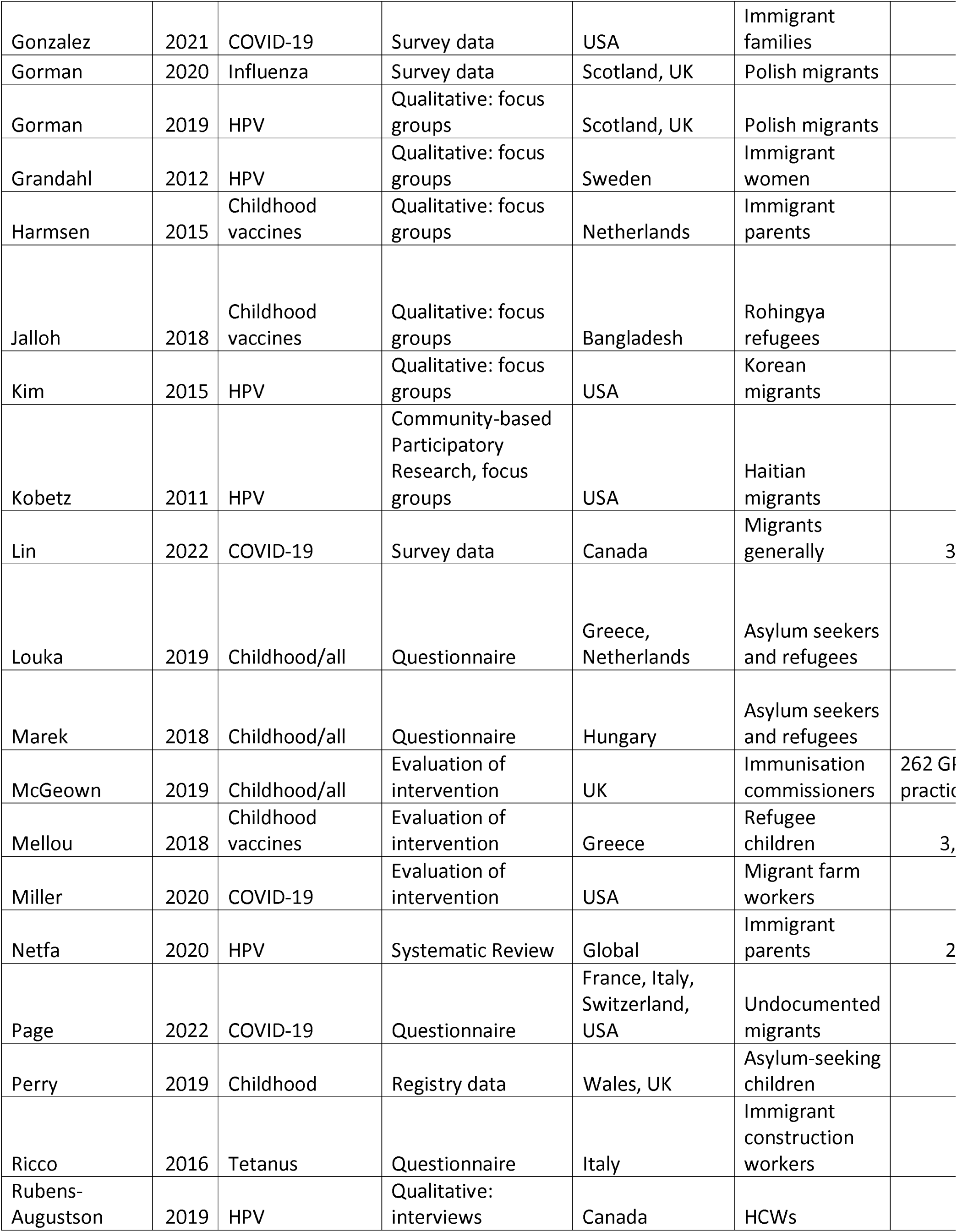
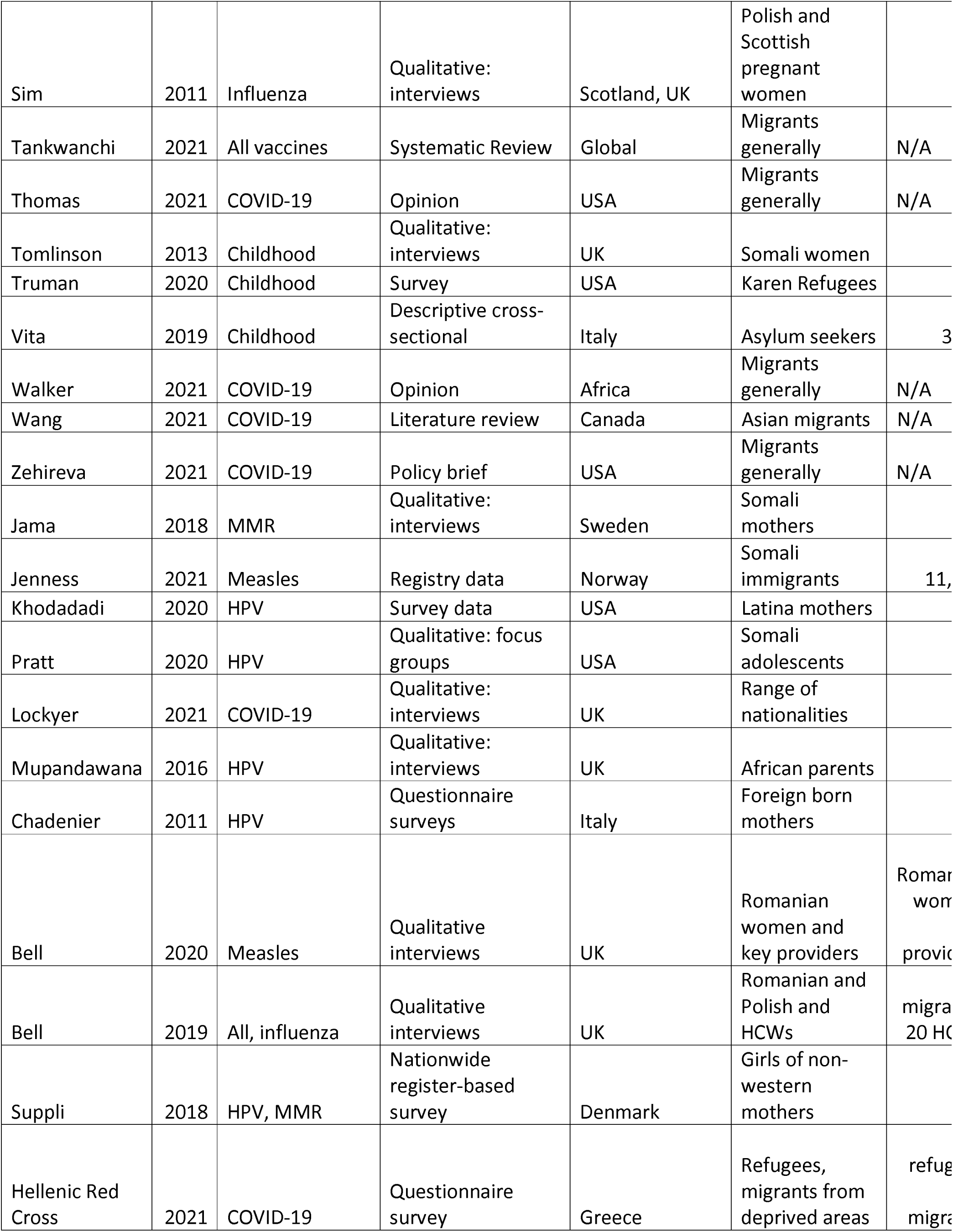
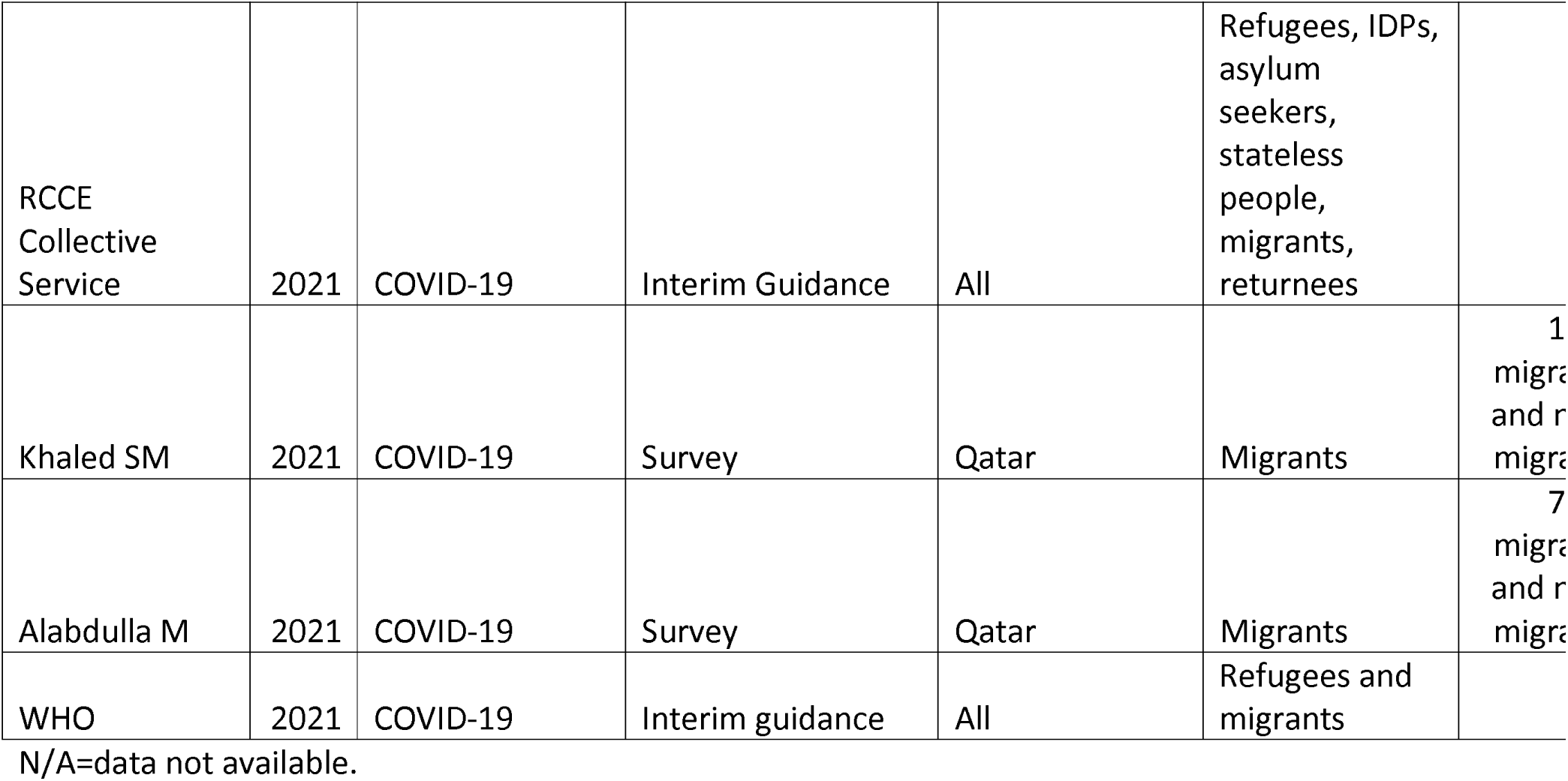
Characteristics of included literature (n=63)

### Personal Factors: What people think and feel about vaccines

Personal confidence in vaccines, such as concerns about safety and side effects are often cited as key drivers of vaccine hesitancy among refugees and migrants (39–45), for example, a study of 1,037 Syrian refugees in Lebanon found that COVID-19 vaccine refusal was significantly associated with negative perceptions about vaccine safety (42). A Canadian study showed that among those who were vaccine-hesitant, a significantly higher percentage of migrants reported concerns about vaccine safety (71.3% vs. 49.5% Canadian-born), side effects (66.4% vs 47.3%) and mistrust in vaccinations (12.5% vs 6.6%) as reasons for vaccine refusal (45). Risk perception has been shown to be important among migrant communities when making vaccination decisions (46), with the perceived dangers of vaccination weighed against complacency around either the need for vaccination (47) or perceptions of danger relating to the specific vaccine-preventable disease (48). In some communities, preferences exist for ‘more natural’ options such as herbal remedies, avoiding human contact, reliance on our immune system (40, 44). A study looking at influenza vaccine attitudes among Polish migrants in the UK found that participant complacency about flu risk was often weighed against trust in the healthcare system and trust in the vaccine (in terms of side effects) when making vaccination decisions (48). One study among refugees and migrants in deprived areas in Greece found that many (111 of 447) did not know whether the vaccine was effective in ending the pandemic, and 91 of 447 respondents did not think it was(49), and despite having various concerns about the COVID-19 vaccine specifically (side-effects, low perception of risk etc) were generally supporting of vaccination more widely (314 of 447 believed vaccines were important for disease prevention).

Trust is major factor in opinions on vaccination (both trust in vaccination itself, and the wider governance and healthcare system of the host country) (31, 43, 48, 50–52). In one UK study among Romanian women around measles vaccines, the active decline in vaccination was linked to distrust in healthcare services, which were partly rooted in negative experiences of healthcare in Romania and the UK(53). Research suggests that low trust among some refugee and migrant populations towards vaccination itself or vaccination systems could be solved by engagement (46, 54–57) with community/religious leaders, relevant NGOs and community groups etc (58). In-depth research should be done in advance to identify and engage with relevant actors, who should be chosen dependent on context (11).

Data also suggest that an individual’s awareness and access to information, which are often dependant on their health and digital literacy, are important factors in vaccine hesitancy. For example, a systematic review on attitudes towards HPV vaccination in migrants found that attitudes often changed for the better once information was given (59), and low awareness about a vaccine or vaccine-preventable disease has been shown to be a key barrier to vaccination (60). Some refugee and migrant populations face specific barriers to information access, such as digital literacy or lack of technology (40, 61), language barriers (11, 32, 39, 40, 47, 51, 61–64), poor doctor-patient communication (41, 60, 64, 65), or a lack of information in an accessible and acceptable format (44, 47, 60, 61, 64), leading to low awareness. The evidence suggests that prior to a vaccination campaign, research should be done on what information is felt to be important among different groups (41, 42, 47, 61, 63, 66). For example, many groups consider safety and efficacy data essential (40, 67). Multiple formats should always be used to increase reach, and these should depend on the context (40). For example, a UK study reported that health visitors felt translated leaflets were not sufficient, as many refugees and migrants struggle with literacy (68). Kim et al noted that, contrary to other studies of vaccination knowledge among US-based immigrants, Korean immigrants in the US often did not receive or understand information disseminated through traditional media such as TV advertisements or radio (69). This shows the importance of information formats, distribution and languages being culturally appropriate and tailored to the target group (50, 66) based on local evidence and informed/co-designed by collaborations with local actors (11, 40, 41, 44, 47, 56, 58, 60). Language barriers are often a problem among refugee and migrant groups, despite increased efforts in some health systems to combat this. In five countries (Australia, Canada, Japan, South Korea, USA), which all ran translated campaigns on COVID-19 vaccination, a lack of details in translated campaigns, inadequacy in the diversity of languages covered and delays in producing translated campaigns were consistently criticized as major hurdles in vaccinating refugees and migrants (70). Multiple languages should be used to disseminate vaccination information, both in written and other formats e.g. interpreters present either virtually or in person to answer questions (11, 50, 63, 64, 70, 71). Six included papers suggest that education programmes should be created for refugee and migrant communities in collaboration with trusted, relevant actors to ensure cultural acceptability and reach (57, 64, 69, 71, 72).

Table 3 summarises factors contributing to low acceptability of vaccination, and some proposed solutions arising from the research.

**Table 3.** Factors contributing to low acceptability of vaccination, and proposed solutions

|  | **Vaccine** | **Population** | **Factors contributing to low acceptability** | **Solutions, strategies, best practices proposed** |
| --- | --- | --- | --- | --- |
| Andrade, UAE | COVID-19 | Migrants generally | Alienation from society, perceived ethnic discrimination, education levels | Educational campaigns, vaccine mandates, increased social integration |
| Aragones, USA | HPV | Latino immigrants | Lack of provider recommendation, fear of side effects | Need for tailored information, preferably from HCWs |
| Artiga, USA | COVID-19 | Migrants generally | Fear of side effects; language barriers to information; fear of links to immigration services | Tailored outreach and education; culturally appropriate information in range of languages; pop-up services to counter lack of trust |
| Berardi, multiple countries | COVID-19 | Migrants generally | lack of details and language diversity in translated campaigns, requirement for undocumented migrants to register, exclusion from healthcare/welfare systems, stigmatisation, | effective community engagement for bidirectional communication and dialog |
| Campeau, USA | Measles | Somali parents | Migration history, and structural marginalisation (which make it hard to trust medical providers) during resettlement, concerns about non-inclusive clinical research, beliefs in immunity as flexible and personalised (developed through encounters with germs, not through vaccination) influence vaccination decisions | Collective social and institutional change and resource redistribution |
| Crawshaw, UK | COVID-19 | Migrants generally | Doctor-patient relationships; language barriers; religious and cultural beliefs; structural inequities; susceptibility to misinformation; conflicting recommendations from country of origin; lack of inclusiveness in vaccination programmes | Participatory approaches, engagement and co-design of vaccination programs/interventions; provide platforms for concerns to be shared without judgement; build trust through transparency |
| Dailey, USA | HPV | Somali parents | Fears of side effects, cultural beliefs, complacency about need for vaccine, lack of information on efficacy | Tailored, culturally appropriate communication needed with stress on health benefits of vaccination, oral information given by a GP/HCW important |
| Deal, UK | COVID-19 | Refugees, asylum seekers, undocumented migrants | Fears of side effects, misinformation circulating, lack of trust in authorities, fears over immigration checks, preference for 'natural' solutions, structural inequity, religious beliefs | Campaigns to increase trust in primary care and educate HCWs about migrant health needs; walk-in/pop-up vaccination centres in trusted places; tailored information in range of languages; avoid stigmatisation; collaboration with community/religious groups or non-governmental organisations |
| El Salibi, Lebanon | COVID-19 | Syrian refugees | Fears of side effects or low vaccine safety, newness of vaccine, COVID-19 vaccine not essential, newness of vaccine, lack of confidence in vaccine | Information campaigns to counter misinformation, collaboration with community leaders, organisations and other key influencers |
|  |  |  | efficacy, preference for 'natural' prevention methods (e.g. social distancing), lack of trust in the system | to re-establish trust |
| Ganczak, Poland | Childhood | Ukrainian | Difficulty finding trusted information on vaccines, low trust in the system and vaccines in home country, misinformation circulating, fears around vaccine safety | Information on vaccination importance and safety to be given by HCWs, training needed to allow HCWs to do this accurately, translated information/material required |
| Gehlbach, USA | COVID-19 | Latinx farm workers | Distrust in the system and medicine, misinformation circulating, financial and social insecurity and inequality, lack of access to internet or other information sources, language barriers | Education in digital literacy; normalisation of vaccination; engage community in decision making around service delivery and outreach; tailored information campaigns |
| Riccardo, EU | Childhood/ all | Migrants generally | Lack of trust in authorities, cultural acceptability and fear of immunisation | Advocate for decreasing inequalities in healthcare, involve and empower community health leaders; use cultural mediators; guidelines training and tools should be created for migrant sensitive advocacy and communication |
| Godoy-Ramirez, Sweden | Childhood | Undocumented migrants | Low trust in system; fear of side effects; instability meaning vaccination becomes a lower priority; previous bad experiences of host | Education for HCWs on migrant health needs, efforts to restore trust in health system; greater focus on tailoring appropriate |
|  |  |  | healthcare system | strategies to specific groups; interventions should be multi-component and dialogue-based |
| Gonzalez, USA | COVID-19 | Immigrant families | Fears of side effects, unsure about vaccine efficacy or feel vaccination is not needed | Immigrants more likely than non-immigrants to trust local public health officials; use elected officials and religious leaders for vaccine information; culturally appropriate, tailored information of vaccine safety, efficacy and access points needed |
| Gorman, UK | Influenza | Polish migrants | Fear of side effects, influence of diaspora/home media; complacency around rare disease; distrust in new vaccines | Promote vaccination on social media through community specific influencers; recruit HCWs from specific community |
| Gorman, UK | HPV | Polish migrants | Influence of distrust in home healthcare system; difficulty understanding UK health and vaccination system; communication and language barriers | Educate parents and HCWs, tailor information and ensure it is accessible and understandable; collaboration with community members |
| Jalloh, Bangladesh | Childhood | Rohingya refugees | Fear of side effects, preference for traditional treatments; religious beliefs; past experiences of vaccination in the camp; fears that 'white' humanitarian workers had ulterior motives behind vaccination | Information campaigns in religious or community meetings; collaboration with community and religious leaders; increased awareness and accommodation of religious or cultural barriers among HCWs |
| Kim, USA | HPV | Korean migrants | Worries about vaccine safety, peer opinions on vaccination, | Recommendation by HCW or other authority (e.g. school) important, school-based education recommended or education in any other existing, familiar environment (e.g. church) |
| Kobetz, USA | HPV | Haitian migrants | Scepticism about vaccine efficacy; fear of side effects and safety; ambivalence; fears about newness of vaccine | Recommendation by doctor important; information and education about side effects and efficacy delivered through trusted sources e.g. community HCW |
| Lin, Canada | COVID-19 | Migrants generally | Fear of side effects, vaccine safety and mistrust in vaccination generally | Pro-active health communication, pop-up COVID-19 vaccine clinics via community- and faith-based organizations |
| Louka, Greece, Netherlands | Childhood/ all | Asylum seekers | Vaccination not always considered important, particularly in country of resettlement (i.e. Netherlands) compared with transit countries (i.e. Greece) | Point of entry to Europe considered best timing for vaccination by asylum seekers; public healthcare system preferred as access point over NGOs; promotional work on vaccination important |
| Netfa, Global | HPV | Immigrant parents | Side effect and vaccine safety concerns; religious and cultural norms; feeling vaccination is not important or needed; distrust in medical provider/pharmaceutical companies | Negative attitudes often changed when information given; information in multiple languages; personal counselling in clinics; educational programmes |
| Page, multiple countries | COVID-19 | Undocumented migrants | Use of social media or community networks as vaccination information source | Community engagement and messaging in multiple languages |
| Perry, UK | Childhood | Asylum-seeking children | Lack of trust; language barriers with HCWs | None given |
| Ricco, Italy | Tetanus | Immigrant construction? Workers | Foreign-born more likely to see recommendation by local public health services as a reason to get vaccinated; more like to state religious/personal belief as a reason to not | Foreign-born workers mostly received doses through occupational health teams |
| Sim, UK | Influenza | Polish pregnant women | Complacency about seriousness of vaccine-preventable diseases; fears around vaccine safety; concerns about being used as 'guinea pigs' | Provide translated, accessible information for range of literacy skills, assistance accessing face-to-face advice |
| Thomas, USA | COVID-19 | Migrants generally | Lack of investment in community partnerships | Integrate community groups and individuals into vaccination processes; including at leadership and decision-making levels |
| Tomlinson, UK | Childhood | Somali women | Risk perception: disease vs vaccine safety, fears of side effects, misinformation, peer opinions, religious concerns especially worry around gelatine content | Oral information from a Somali-speaker; develop closer relationships between health providers and community; work with religious leaders |
| Truman, USA | Childhood/all | Karen Refugees | Longer time spent in US associated with more reluctance to vaccinate | Culturally fit interventions and education programmes recommended |
| Walker, Africa | COVID-19 | Migrants generally | Fears around hidden agendas of 'the west', misinformation, different policies about vaccination across countries (e.g. Tanzania, Madagascar) | Inclusion of community groups, religious leaders etc; sharing information online in migrant languages |
| Wang, Canada | COVID-19 | Asian migrants | Misinformation on social media; language and literacy barriers; mistrust in healthcare services; hesitancy among HCWs; systematic racism and mistrust | Incorporate cultural competency in healthcare; tailored information for preferred languages; directly address specific concerns; HCWs from migrant communities can aid communication |
| Jama, Sweden | MMR | Somali mothers | Fear of side effects (autism); bad previous experiences with HCWs; misinformation from word-of-mouth; stigmatisation and feeling their views are not listened to by HCWs; peer opinions on vaccination | Interventions that focus on communication mechanisms; particularly through nurses |
| Jenness, Norway | Measles | Somali immigrants | More time in host country and urban location associated with lower uptake | Tailored strategies for community; social network analysis needed to identify influencers in communities to collaborate with; improved communication |
| Khodadadi, USA | HPV | Latina mothers | Low perceived risk of HPV associated with reluctance to vaccinate | Education and improvement of health literacy to increase risk awareness |
| Pratt, USA | HPV | Somali adolescents | Scepticism or fear about vaccines in general; religious and cultural norms; |  |
|  |  |  | feelings of stigmatisation |  |
| Lockyer, UK | COVID-19 | Mix | Fear of side effects and vaccine safety; structural inequality and precarity, confusion and distrust in authorities and traditional media; conflicting or negative information from home country or social media, existing distrust in institutions | Systematic monitoring of misinformation on social media and responding sensitively; inform HCWs about circulating misinformation; harnessing connections with trusted community network; providing information in multiple languages |
| Grandahl, Sweden | HPV | Immigrant women | Communication barriers; cultural health norms, fears of side effects of low efficacy | Ensure availability of interpreter; more information required; translated invitation letters |
| Mupandawana, UK | HPV | African parents | Cultural and religious norms, complacency about VPD risk; risk perception, preferring natural prevention methods (abstinence); side effects, misinformation; distrust of 'the West' | Tailored information that addresses cultural and religious concerns; create smaller subgroups for targeted communication; video/story-based educational material, collaboration with religious or other leaders |
| Rubens-Augustson, Canada | HPV | HCWs | Language barriers; lack of appropriate information resources; cultural and religious factors; limited HCW time | Targeted health promotion eg, in schools, ensuring access to appropriate personnel, culturally sensitive risk communication |
| Tankwanchi, Global | All | Migrants generally | Fears and misinformation about safety; limited knowledge of VPDs and vaccines; distrust of host health | Tailored, community-based immunisation service delivery with migrant-friendly health systems and policies that affirm |
|  |  |  | systems; language barriers; religious beliefs | and protect human rights and dignity |
| Harmsen, Netherlands | All | Immigrant parents | Lack of time/information given to patients by HCWs; fear of side effects; language barriers; newness of vaccines | Information in own language; more oral information from HCWs or in specific educative meetings |
| Bell, UK | All | Polish and Romanian migrants | Expectations largely built on knowledge and experience from Poland and Romania; greater refusal of influenza vaccine due to perceptions around lack of efficacy | HCWs to explain how the health system works and clarify expectations; outreach to those facing barriers to healthcare; translated information; use pictograms or pictures; improve access to interpreting and translation services; use views and expectations of service users to shape services |
| Bell, UK | Measles | Romanian women and key providers | Concerns around vaccine safety; distrust in healthcare services, which were partly rooted in negative experiences of healthcare in Romania and the UK | Tackle cultural and linguistic barriers; strengthen provider-service user relationships; establish trust providers must find ways to connect with and develop a greater understanding of the communities they serve |
| Hellenic Red Cross | COVID-19 | Refugees and migrants in deprived areas | Don't think vaccines are effective in ending the pandemic; didn't consider themselves at risk; side-effects; not | Creation of information material with Q&A type information in all spoken languages; because respondents |
|  |  |  | adequately informed about the vaccine; did not believe COVID-19 existed | sourced information from the internet, provide robust sources of internet information; use a combination of information approaches – printed material, posters, community meetings; establish a COVID-19 handbook for all involved in vaccine delivery |

HCW=Healthcare worker. GP=General practitioner.

**Table 4:**
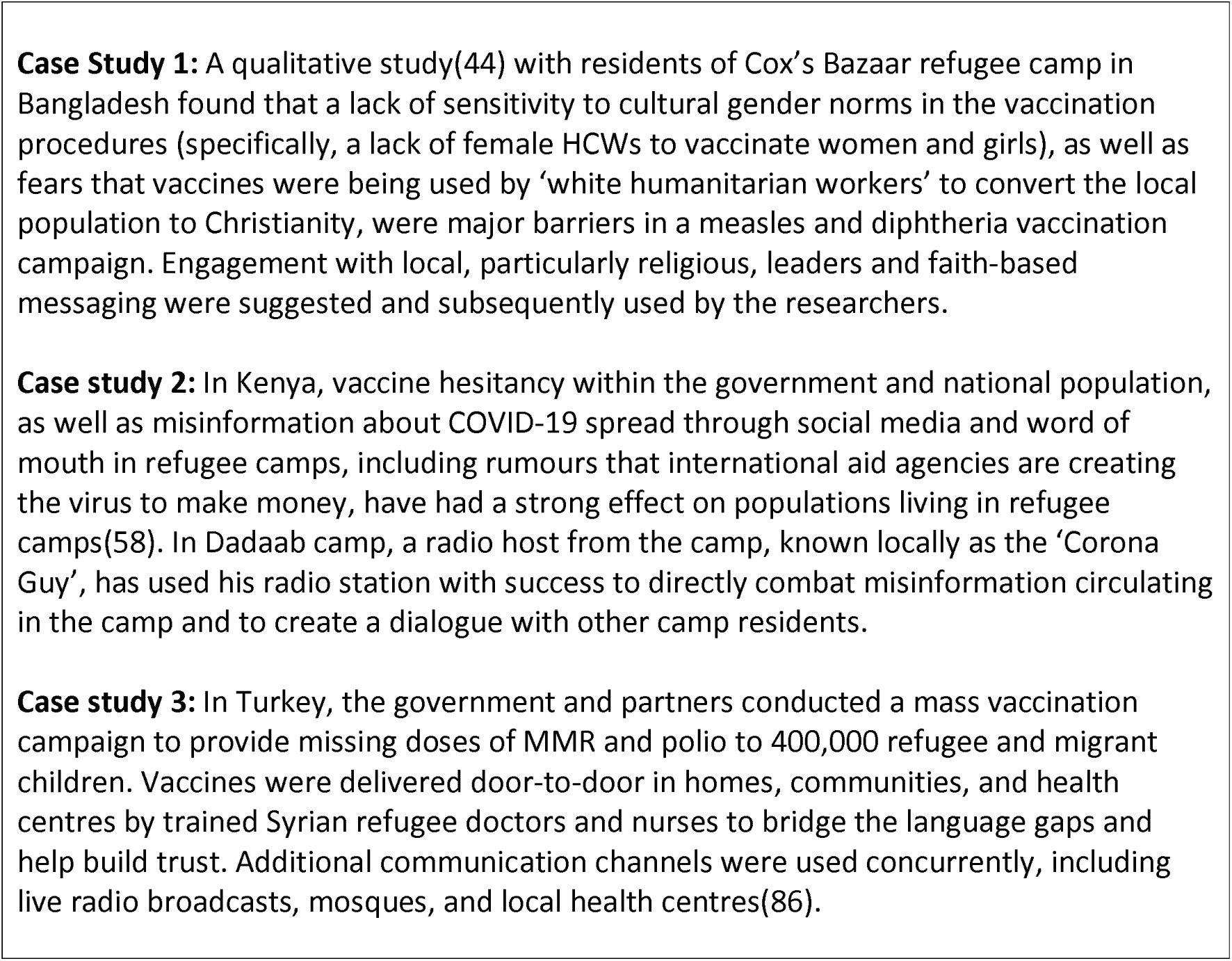
Case studies for increasing vaccine acceptance in humanitarian settings

### Social processes: drivers or inhibitors of individual motivation to seek vaccination

Included research strongly suggests that the perceived acceptability of vaccines among refugee and migrant groups is highly dependent on context and social processes, including historical, economic, religious or political factors in different countries or regions (31). In some cases, migrant expectations may be built on knowledge and experience from their home countries (32). For example, a lack of trust in vaccines in a refugee and migrant’s country of origin, or the influence of factors from the home country or diaspora media can be an important factor in vaccine confidence in some groups (34, 39, 47, 73). However, distrust of vaccination can also originate or worsen after arrival in the host country; for example, a study in the USA showed that amongst Karen refugees, a longer time spent in USA was associated with lower levels of perception that vaccinations were safe (72). Another study from Norway found that children born to mothers residing in Norway for more than 6 years had lower measles coverage compared to those residing less than two years prior to their birth, with coverage overall declining between 2000 and 2016 (74). This could be due to social exclusion or alienation after arrival (31, 75) or precarity in resettlement (76), which have both been shown to negatively affect vaccine confidence in included papers. A study in the UK found that the more confused, distressed and mistrusting the participants felt during the COVID-19 pandemic, the more likely they were to be hesitant about uptake of the COVID-19 vaccine (39).

Political and economic processes can also be important factors affecting motivation to vaccinate, as shown in two studies from Qatar, which found that migrant status was associated with lower levels of vaccine hesitancy and reluctance to accept a vaccine compared with Qatari nationals. The authors noted that migrants’ residency status in Qatar is tied to employment contracts, suggesting they will be more accepting of government or employer policy(77, 78). Historical and structural racism in the host country can also influence motivation to vaccinate in migrant communities (39, 58, 61, 76), for example, fears that certain ethnic groups or communities may be used as ‘guinea pigs’ in the COVID-19 vaccine roll-out (39, 40, 48, 61), often originating from historical events (e.g, the Tuskegee study). In Africa, fears that there are hidden agendas of ’the West’ behind vaccination campaigns are reported to be circulating, including among refugee and migrant groups (58). Interestingly, the same concerns around vaccines based on distrust in ‘the West’ were brought up by African refugees and migrants living in the UK (79), supporting the theory that home country context and diaspora media remain important factors in the perceived acceptability of vaccination.

Misinformation on vaccines can spread rapidly through social media or word of mouth and may have a strong influence on individual and community vaccine confidence, particularly in communities, such as many refugee and migrant communities, where distrust already exists and/or who have more limited access to robust public health information (39, 40, 61, 80). Questionnaires of undocumented migrants in Paris, Rome, Baltimore and Milan have suggested that using social media or community networks as the preferred source of vaccination information was negatively associated with demand (81). A study in Bradford, UK, showed that rapid local and targeted responses to specific misinformation can be a solution and cited an example of a video produced by Bradford city council in Urdu and Punjabi which debunked a specific conspiracy story spreading in these local communities, which was reported to be effective (39).

Religious norms or expectations are another key social process that may influence migrants’ motivation to vaccinate and have been previously shown affect perceived acceptability of vaccines (41, 44, 82). A study in Italy found immigrant workers more likely than non-immigration workers to state religious belief as a reason to not vaccinate (82); another study of Lebanese immigrants in Australia found that religious values around health played a major part in vaccination decision making (66) and a lack of female vaccinators (with male HCWs vaccinating females seen as unacceptable due to Islamic principle of purdah) was seen as a barrier to vaccination among Rohingya refugees in camps in Bangladesh (44). Collaboration with religious and cultural leaders is important (44) and education, outreach activities, and tailored information campaigns should draw on specific religious values where appropriate for the target community (66). For example, Muslim communities may have concerns about whether vaccinations contain pork (46), which is a crucial factor to be addressed in communications.

Low knowledge of refugees and migrant health needs or of eligibility to healthcare among healthcare workers (HCWs) is another social process that may drive migrant motivation to vaccinate. HCWs were often considered the main expected source of information among refugees and migrants HCWs are often considered the main expected source of information among refugees (40, 64, 83, 84), yet Rubens-Augustson showed in a qualitative interview study in Canada that HCWs working with migrants often feel that communication barriers and inability to address cultural or religious concerns restricted them in informing refugee and migrant patients about HPV vaccines (64). Education programmes for HCWs on working with refugee and migrant populations (56, 85) and on refugee and migrant entitlement to healthcare and vaccination (40, 64, 83, 84) were widely recommended in the literature, with on study showing increased uptake of catch-up vaccination in a centre for unaccompanied asylum seeking children with increased awareness of migrant vaccination needs among HCWs (85).

### Physical considerations: the ability of individuals to be reached by, reach or afford recommended vaccines

Included papers described the accessibility of vaccination as potential driver of vaccine hesitancy among refugee and migrant populations, with migrant entitlement to health and vaccination systems in the host country a critical factor (54). In the context of the COVID-19 vaccine roll-out, it is as yet unclear in several countries the extent to which refugees and migrants will have access to vaccines based on migrant status, with particular concerns around undocumented migrants accessing health services and the vaccine at present in several European countries where coverage is relatively high (4).

Evidence suggests groups such as undocumented migrants residing in high-income countries may have fears about data sharing, immigration checks, lack of eligibility and other immigration-related concerns, highlighting that it is essential going forward for personal data collection associated with vaccination campaigns to be kept minimal and for undocumented migrants and for these groups to be offered access points where they feel safe from immigration enforcement (40, 43, 55, 61, 86). A study among migrants living in informal settlements in Italy showed that irregular migrants had the lowest COVID-19 vaccine coverage 15.7%) of all migrant groups (asylum seekers 28.9%, other residence permit 38.5%) (52). A need to create awareness among migrant population, as well as HCWs, about their entitlement was reported (40, 43, 56); perceived ineligibility for vaccination and/or free healthcare in the host country can contribute to hesitancy (40). Papers highlight several examples of good practice specifically around undocumented migrants and addressing barriers to vaccination: for example some governments have removed healthcare entitlement barriers to testing and vaccination for COVID-19 or stated that vaccines will be available irrespective of residence status, and can be acquired anonymously with no links to immigration enforcement and the Colombian government has provided a 10-year temporary protection status to Venezuelan migrants which will allow them to register for vaccination (4, 11, 87).

Convenience of access points is often a key factor in vaccination decision-making among refugees and migrants, particularly those for whom losing a day of work to visit a distant vaccination centre may entail significant financial loss. Specific access points should be created, advised by and in collaboration with local actors (55); for example, a US-based policy brief has recommended that legislation is passed to allow pharmacists to administer vaccines in underserved communities including migrant communities (88). Interventions which ‘take vaccination to migrants’ rather than expect migrants to present themselves for vaccines have historically had success: in Italy, migrant construction workers mostly received tetanus and diphtheria vaccines through occupational health as opposed to community-based primary care (82) and a door-to-door vaccination programme for refugees in Greece in collaboration with local non-governmental organisations saw >20,000 childhood vaccination doses delivered (89). Good vaccine uptake was noted during an intervention offering on-farm COVID-19 vaccine to migrant farmworkers in the USA (90) and vaccination uptake was observed to be much higher in a migrant reception centre in Italy when doctors came to centre and gave vaccines when migrants arrived (91). In a paper by Chauhan et al, proactive contacting of migrants for their vaccinations was suggested to maximise uptake (85) and Thomas et al suggest proactivity in bringing vaccination to migrants rather than expecting them to present at vaccination services improved uptake (50). One of six recommendations suggested in one paper to increase uptake in migrants was to create a reminder system for specific vaccines in primary care, so that patients can be reminded of and offered their missing vaccinations when presenting for other reasons (64). One study from Denmark found that migrant girls were more likely than Danish-born girls to have HPV after receiving a reminder (92); on the other hand a study around measles vaccination in Romanian migrants in the UK found that the blanket approach of sending text message reminders appeared to be not that effective, due to language and literacy barriers (53). The mobility of some migrant populations may need to be considered especially when multiple doses of vaccines are required (43, 93), which may be an important issue in COVID-19 vaccine roll out with most vaccines requiring two doses. A study among migrants living in informal settlements in Italy found that those who were in transit had significantly lower COVID-19 vaccine coverage (52). Population mobility in Greek refugee camps was overcome by door-to-door visits by staff from non-governmental organisations to actively record vaccination status within two weeks of each vaccination intervention (89).

Several papers suggest that economic barriers and affordability of vaccines are also important in decision making for some refugees and migrants (40, 47, 52, 53, 57, 62, 64, 67, 94). This can include direct costs; for example, Louka et al found that asylum seekers in Greece and the Netherlands became less likely to accept vaccination as the cost increased (57). In another report, discussing HPV vaccination in Canada, healthcare providers recommended publicly funding vaccination as a key facilitator to maximise uptake among refugees and migrants (64); in a qualitative study in the USA, the majority of refugees and migrants interviewed responded positively when asked about HPV vaccine intent provided the vaccine be affordable. In Italy foreign-born mothers were reported to be less likely to be willing or able to pay 100 Euros for their daughters to be vaccinated(62). In addition, a lack of clarity about charges for vaccination and/or healthcare may put migrant off seeking vaccination(53). Costs and other factors may influence where migrants choose to get vaccinated, with one UK study reporting Polish migrants returning home for cheaper vaccines (chicken pox) and/or after the birth of a child for routine vaccines, with numerous implications at a service provider level(32). Indirect costs, such as travel costs, and wages lost from time off work are also an important factor, particularly for groups with precarious status or working in low-skilled jobs. One paper on vaccination among Latinx immigrants in the US suggested that workers from this group may not be given permission or time off to attend a COVID-19 vaccination appointment, and be unable to pay for transportation to distant vaccination sites, recommending that vaccination campaigns should educate employers to give employees time off for vaccination as well as ensuring that vaccination sites are easily accessible to underserved communities (86).

Figure 2 summarises key behavioural and social drivers of vaccination, and solutions and strategies to tackling it in the context of COVID-19 roll-out, compiled from the included literature in this review.

**Figure 2.**
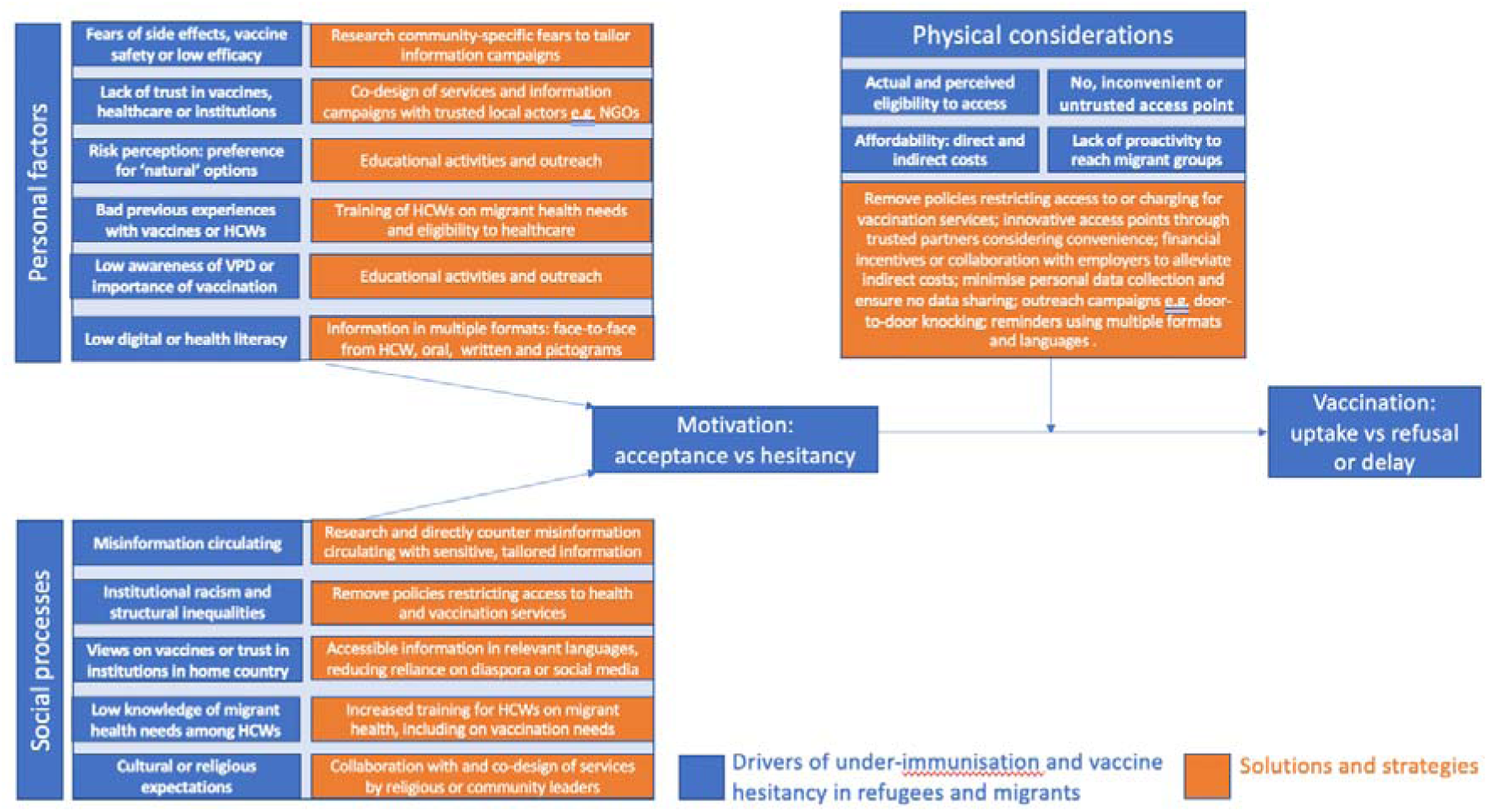
Solutions and strategies for increasing COVID-19 vaccination uptake in refugee and migrant populations, based on the Increasing Vaccination Model (20–22)

## Discussion

This review has compiled a wide range of literature pertaining to drivers of under-immunisation and vaccine hesitancy in refugee and migrant populations, and solutions and strategies to addressing it. We found a stark lack of research from low- and middle-income countries and humanitarian contexts, a situation that needs to be urgently rectified as COVID-19 vaccine roll-out gathers pace beyond high-income countries. We found that there are a range of factors driving under-immunisation and hesitancy in some refugee and migrant groups, with the acceptability of vaccination often deeply rooted in social and historical context and influenced by personal risk perception. Unique issues relating to awareness and access to vaccination for some refugee and migrant populations also influences vaccine motivation and needs to be better considered in research, policy making, and service delivery in the context of COVID-19 vaccination. There is an urgent need for more robust research on the influence of circulating misinformation on social media platforms, and the impact of information from other sources (eg, diaspora media) on COVID-19 vaccine uptake in refugees and migrants. Lessons learned to date suggest that there are relatively simple solutions and strategies to improve COVID-19 vaccine uptake in refugees and migrants that will be context-dependent, with a focus needed on meaningful community engagement, patient/provider interactions and building trust, strong risk communications, designing and delivering tailored information that is context and audience specific, and identifying innovative migrant-friendly access points.

Drivers of under-immunisation and vaccine hesitancy in refugee and migrant populations are complex, multi-factorial and highly context-dependant, and it is not possible from the evidence we have to explore similarities and differences between these diverse settings and regions of the world. Solutions and strategies are also not necessarily generalisable between countries, regions or migrant groups and few are ever robustly tested to explore their effectiveness on actually increasing vaccine uptake. Important contextual factors include economics and politics in the host country, particularly related to healthcare access, the precarity of specific migrant groups, cultural and religious norms around healthcare and vaccination, historical factors or structural racism in the host country as well as misinformation or vaccination norms from migrants’ home countries. The WHO’s Tailoring Immunization Programmes (TIP) framework (95) and more recent Data for Action survey tools for COVID-19 (22), aim to support countries to assess and monitor vaccine hesitancy and low uptake in susceptible populations, recognising that behaviours are complex, populations are diverse ,and that a ‘one size fits all’ approach will not work. These approaches stress the importance of doing robust research prior to any vaccination campaigns to identify key behavioural and social drivers of vaccination to vaccination and innovative strategies. Working to better understand how drivers of under-immunisation and vaccine hesitancy in refugees and migrants impacts on vaccine uptake, as well as working more closely with affected communities, will ensure effective solutions are developed and delivered. Practical approaches to strengthening demand and uptake in refugees and migrants for COVID-19 vaccines has been outlined in a recent WHO Operational Guide (https://www.who.int/publications/i/item/WHO-2019-nCoV-immunization-demand_planning-refugees_and_migrants-2022.1)

Our work has highlighted the importance of social processes and physical considerations in influencing vaccination motivation in some refugee and migrant populations, and that these processes are often unique to refugee and migrant populations – for example a lack of entitlement to access the health and vaccination system in the host country, or lack of trust in institutions and institutional racism and other structural barriers. We have found that while concerns around vaccination or misinformation may travel with refugees and migrants from their home countries, or circulate in diaspora media or social groups, studies have also shown that refugees and migrants residing in the host country for longer are more likely to be hesitant towards vaccination, suggesting their hesitancy originates after migration due to precarity, structural inequalities or social exclusion in the host country. Cultural or religious norms were often also cited in included literature as important drivers of under-immunisation and vaccine hesitancy in refugee and migrant populations, especially where these norms are not shared by the host country or host country health system. Key to tackling this is for countries to explore innovative refugee-. migrant-friendly access points for distribution of COVID-19 vaccines and ensure HCWs and vaccine deliverers are trained about the unique needs of refugees and migrants (96).

We have shown that personal factors affecting an individuals’ motivation to vaccinate often revolve around risk perception in refugee and migrant groups, with the perceived safety of the vaccine in question and trust in institutions and healthcare services, which is often based on previous experience. This is much in line with drivers of hesitancy in most populations, with risk perception based on concerns around vaccine safety and side effects usually stated as key factors in COVID-19 vaccine hesitancy generally (97). An individuals’ awareness of the vaccine, the risk associated with the specific vaccine-preventable disease and the importance of vaccination generally are also key factors affecting motivation, with low knowledge of any of these often associated with hesitancy. Multiple solutions were proposed in the included literature, often revolving around educational activities with specific community groups, co-design, and outreach and tailored information campaigns in a range of formats and languages. A recent RCCE interim-guidance report on risk communication and community engagement for COVID-19 vaccines in marginalised groups stresses that advanced planning takes place to identify barriers to COVID-19 vaccines (considering gender and intersectoral needs, amongst others) and that it is essential that all new initiatives place community engagement at each point of the process because perceptions and information will change, and that preferred and trusted communication channels that meet a range of different communication needs are found and used (98).

Greater consideration too must be given to new and rapidly evolving drivers of vaccine hesitancy globally, including the influence of social media-based communication as a major source of vaccine misinformation (99). One COVID-19 survey among refugees and migrants in Greece found that 275 of 447 respondents said their main source of information about COVID-19 disease was through social media (49). WHO has drawn attention to the challenge of the ‘infodemic’ or misinformation and disinformation pandemic in the context of COVID-19, calling for universal access to credible health information and efforts to tackle these important new challenges. Specific misinformation should be directly addressed in communities where it is known to be circulating, using sympathetic and transparent messaging in a range of formats and languages. The IFRC has recently developed a series of resources to tackle rumours and misinformation circulating in communities, an information pack on effectively listening and responding to communities around COVID-19 and ensuring feedback mechanisms are in place, including a survey to gain specific data at a community level on individual’s perceptions of COVID-19 vaccination and barriers to access(100).

These findings hold direct relevance to current efforts to ensure high levels of global vaccine coverage for COVID-19 vaccines and highlight the urgent need for a concerted international effort to understand, analyse and overcome vaccine hesitancy. Several guidance documents provide policy actions for strengthening delivery and uptake of COVID-19 vaccines in refugees and migrants, and they are summarised in Table 5. Further research is warranted that places greater emphasis on better understanding vaccination motivation and barriers to vaccination in refugees and migrants, and robustly tests strategies and solutions to better understand their effectiveness in increasing uptake (Table 6). Renewed efforts and investment must be placed on supporting countries to collect, analyse, and source disaggregated data pertaining to vaccination and migration. The near complete absence of vaccine uptake data for COVID-19 vaccines in most high-income countries, with which to inform real-time evidence-based service delivery, is a stark reminder of how just how invisible refugee and migrant populations still are, a situation that now needs to be urgently rectified if we are to improve health outcomes in these groups and meet the regional and global goals of WHO’s new Immunization Agenda 2030 (101). Of key importance now is to ensure marginalised refugee and migrant populations are specifically included in national vaccine-delivery plans of low-middle- and high-income countries, through initiatives including the COVAX Facility and the COVAX Humanitarian Buffer as a last resort, as we advocate for more rapid roll-out of COVID-19 vaccines in low- and middle-income countries and humanitarian settings and promote global vaccine equity.

**Table 5:**
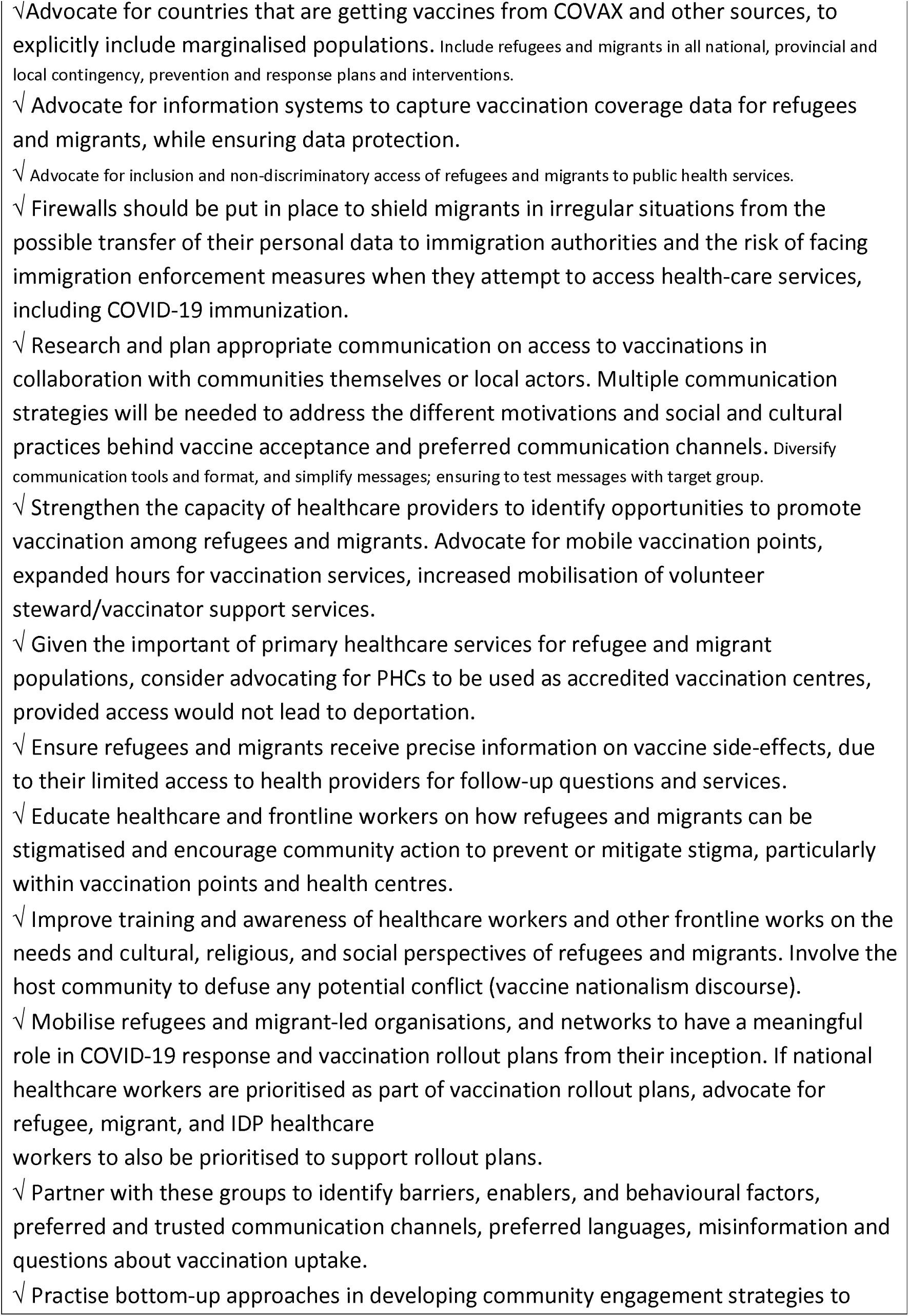
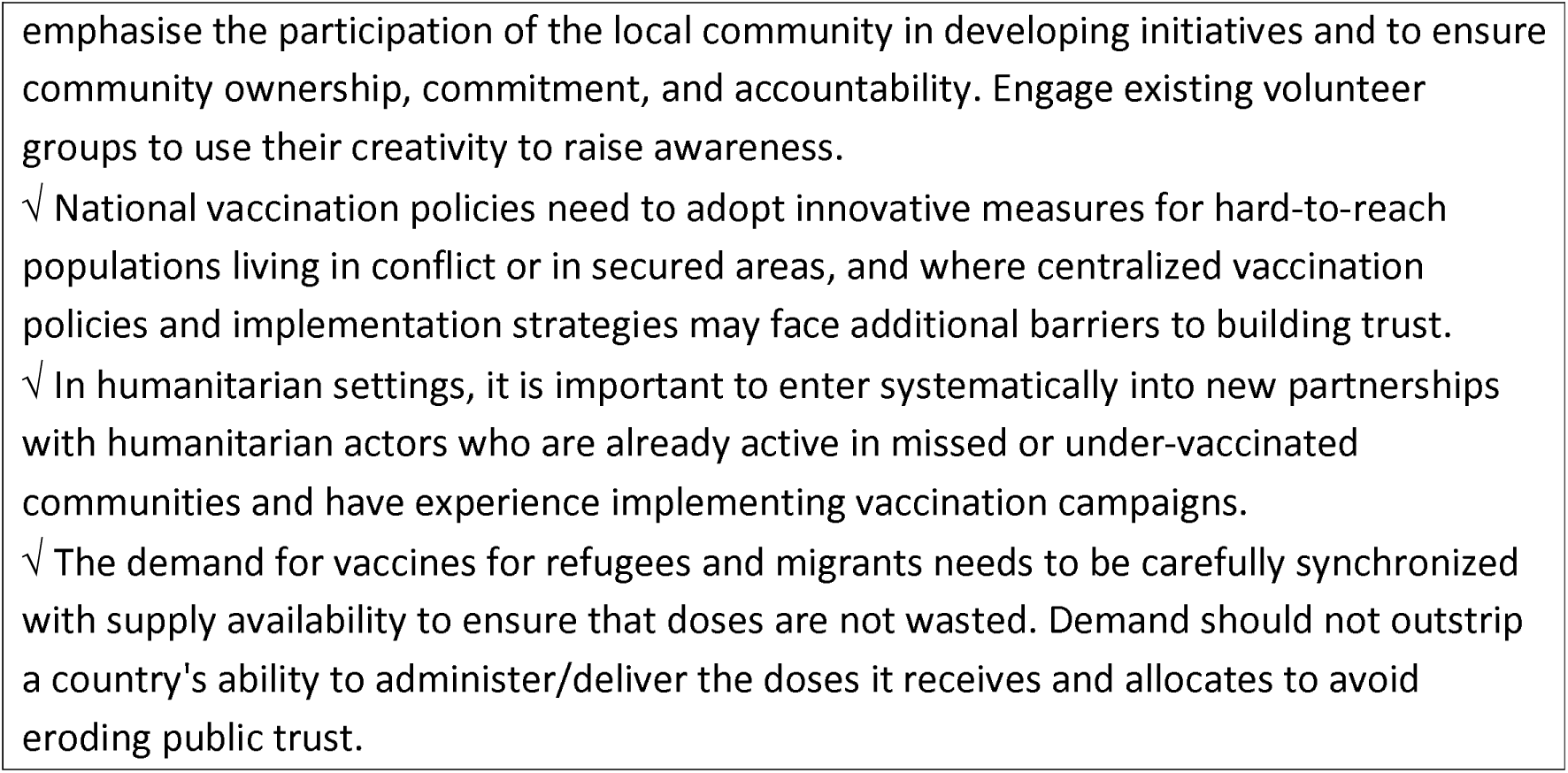
Summary of policy actions to support strategies for COVID-19 vaccine roll out in refugees and migrants (Adapted from (86, 88, 103))

**Table 6:**
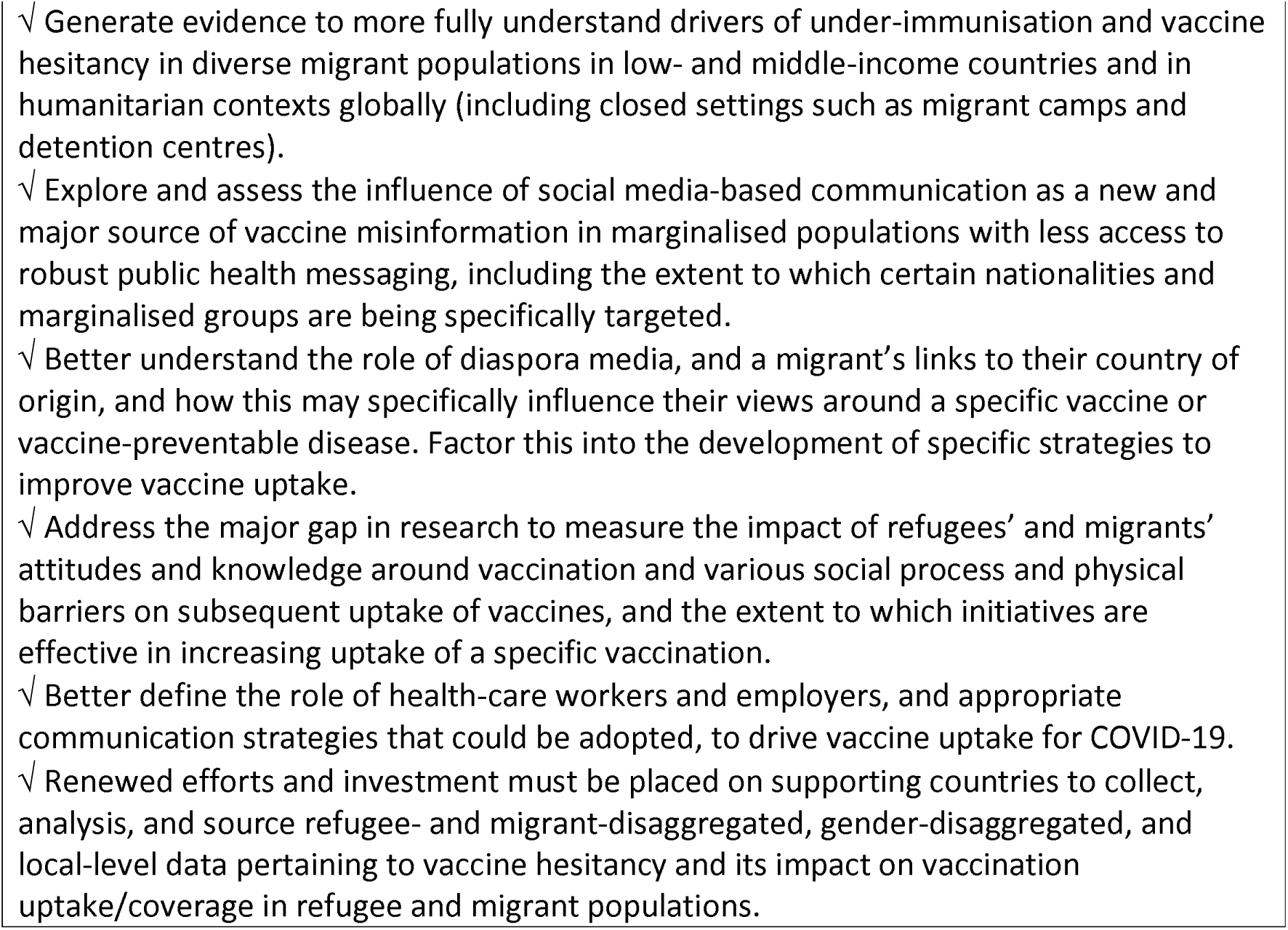
Further research

## Data Availability

All data produced in the present work are contained in the manuscript

